# Estimating the economic burden of typhoid fever in the Democratic Republic of the Congo

**DOI:** 10.64898/2026.09.07.26362464

**Authors:** Yongha Hwang, Robert Lumbala, Jung-Seok Lee, Jules Mbuyamba, Jean-Paul Kumbukama, Mohamadou Siribie, Seung Eun Kyung, Pierre Cédric Khuwa, Narcisse Diakiese, Juvénal Dienda, Hyonjin Jeon, Birkneh Tilahun Tadesse, Marie-France Phoba, Florian Marks, Octavie Lunguya

## Abstract

**Background:** Typhoid fever, despite being preventable, places a significant burden on low- and middle-income countries. Although understanding of the global burden of typhoid fever is improving, evidence on the economic burden of typhoid fever remains limited. This study estimated the cost of illness associated with typhoid fever in the Kisantu Health Zone of the Democratic Republic of the Congo.

**Methods:** A standardized patient-level survey was conducted to estimate the cost of illness associated with typhoid fever. Patients with typhoid fever were recruited from eight health centers and one hospital. Multiple interviews were conducted to capture the full costs incurred throughout the illness episode.

**Results:** From March 2022 to March 2025, among 140 enrolled patients, 91 were classified as mild cases and 49 as severe cases with suspected intestinal perforation/peritonitis. The mean cost of illness for typhoid fever was US$ 108 (95% CI: US$ 90–127) for mild cases and US$ 579 (95% CI: US$ 506–663) for severe cases, indicating that costs for severe cases were more than five times higher than those for mild cases. All direct medical costs were paid out-of-pocket by patients, and no patients were covered by insurance or government subsidies.

**Conclusions:** Typhoid fever imposes a substantial economic burden in the Democratic Republic of the Congo. These findings provide important evidence for evaluating the economic value of typhoid conjugate vaccine introduction in high-burden settings.

## INTRODUCTION

In 2021, typhoid fever caused an estimated 7 million cases and more than 93,000 deaths worldwide(1). Caused by Salmonella enterica serovar Typhi (S. Typhi), typhoid fever continues to pose a substantial public health burden, particularly in low- and middle-income countries (LMICs), despite the availability of effective preventive measures, including improvements in water, sanitation, and vaccination (2, 3). Increasing antimicrobial resistance has reduced the effectiveness of commonly used antibiotics and may increase the likelihood of complications such as intestinal perforation or death (4, 5).

Recent epidemiological studies have substantially improved estimates of global typhoid incidence and mortality (6). In contrast, information on the costs incurred by patients and households remains comparatively scarce (7). Only 13 studies reporting empirical cost data on typhoid fever have been published across 11 countries, four of which were in Africa (Malawi, Niger, Nigeria, and Tanzania). The limited availability of primary cost data constrains economic evaluations of typhoid control strategies, including vaccination.

The World Health Organization (WHO) recommends the use of typhoid conjugate vaccines (TCVs) in endemic settings because of their suitability for young children and longer duration of protection (8). Following Gavi’s financial support of US$ 85 million to support the introduction of TCV in developing countries (9), several countries including Pakistan, Liberia, Nepal, and Zimbabwe have introduced TCV (10, 11). Although TCV has been introduced in several countries, evidence on the economic burden of typhoid and the economic value of implementing TCV campaigns in African settings remains limited (12).

To address the existing knowledge gaps, the European & Developing Countries Clinical Trials Partnership funded a program in 2017 entitled “The Effect of a Novel Typhoid Conjugate Vaccine in Africa: a multicenter study in Ghana and the Democratic Republic of the Congo”. To improve understanding of the economic burden of typhoid, a cost of illness (COI) survey was conducted as part of the project in the Democratic Republic of the Congo (DRC).

The DRC has experienced multiple large-scale typhoid outbreaks due to poor infrastructure and health services (13). A recent population-based surveillance study identified the DRC as the study site with the highest typhoid incidence among participating African countries, exceeding 300 cases per 100,000 population (5). The study also found that school-aged children aged 2 to 15 years were at the highest risk across all study sites. These incidence estimates were substantially higher than the incidence rate threshold of 100 cases per 100,000 people, which the WHO considers as ‘high incidence’ (14). One study carried out in the DRC, estimated 72.2% of *S. Typhi* were co-resistant to certain antibiotics, and an additional 33.3% were multidrug-resistant (4). These findings suggest that typhoid fever remains prevalent and continues to pose a significant public health challenge.

This study aimed to estimate the patient-level cost of illness associated with mild and severe typhoid fever among patients in the Kisantu Health Zone, DRC.

## METHODS

The healthcare structure in the DRC is organized into Health Zones. Fever surveillance was implemented in the Kisantu Health Zone, which has an approximate population of 200,000. Hospital Saint-Luc in Kisantu served as the primary site for the economic evaluation component of the study. Patients of all ages residing in the surveillance area were eligible if they presented with a tympanic temperature ≥38.0°C, an axillary temperature ≥37.5°C, or a history of fever lasting at least three consecutive days within seven days before presentation. In addition, patients suspected of having typhoid-related intestinal perforation or peritonitis requiring surgery (even in the absence of laboratory confirmation) were also eligible.

Typhoid patients were categorized into two groups: mild and severe. A mild case referred to patients with a positive blood culture for *S. Typhi*, while a severe case referred to patients suspected of intestinal perforation/peritonitis due to typhoid fever requiring surgery (even in the absence of laboratory confirmation). Patients who were either laboratory-confirmed as *S. Typhi* or identified as severe cases were recruited to participate in the study and interviewed.

A standardized patient-level survey was implemented across the Kisantu Health Zone. Patients completed up to three follow-up interviews, depending on the duration of their illness (Table 1). Patients with blood culture-confirmed typhoid fever or those identified as severe cases were enrolled in the study. Each interview collected information on patient expenditures and income losses. The second and third interviews were conducted after 30 to 40 days and 90 to 100 days, respectively, if patients had not yet recovered. Interviews were conducted during the period from March 2022 to March 2025.

**Table 1.**
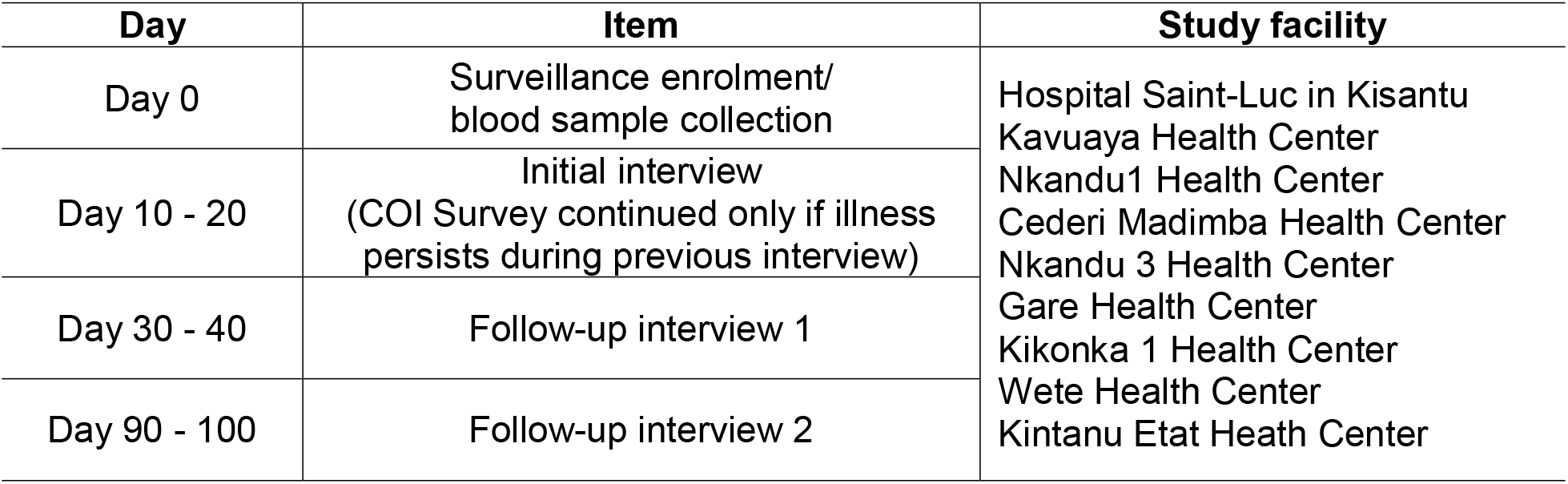
Schedule of follow-up.

The COI survey captured three categories of costs. These were direct medical cost (DMC), direct non-medical cost (DNMC), and indirect cost (IC). The DMC included consultation fees, medications, laboratory tests, and other medical expenses associated with typhoid treatment. In addition to the interview, hospital invoice records were accessed to capture the costs covered by third-party payers, such as insurance or government subsidies. The DNMC included any expenditure associated with meals, accommodation, and transportation for patients and accompanying household members. The IC included productivity losses due to lost wages or missed school days, as well as time losses incurred by caregivers and substitute laborers.

To estimate productivity loss, patients were asked about their daily wage loss. For students who did not earn any wages, productivity loss was approximated using government expenditure per primary student (15, 16). Daily government expenditure per primary student was estimated by dividing annual expenditure per student by the assumed 200 school days per year. If any patients passed away due to typhoid during the study period, the productivity loss from premature death was estimated using the human capital approach (17). Remaining life expectancy was calculated by subtracting age at death from life expectancy from the United Nations Population Division (18), and the resulting years of productivity lost were multiplied by the annual minimum wage and discounted at a 3% annual rate.

In addition to estimating out-of-pocket (OOP) expenditures, non-OOP costs such as governmental or health facility subsidies, or public or private insurance coverage information was assessed. Hospital records for all COI patients were accessed and analyzed.

Bootstrapping was applied to address uncertainty and account for the skewed distribution of cost data. All estimates were presented with 95% confidence intervals. All costs were converted to 2022 US$ using World Bank official exchange rates (19).

### ETHICS STATEMENT

The research protocol for COI was approved by the Institutional Review Boards of the International Vaccine Institute and the School of Public Health at the University of Kinshasa. Written informed consent was obtained from all patients who participated in the COI study. For children aged 12 to 17 years, assents were obtained from the child, and consent was sought from their parents. For children younger than 12 years, consent was obtained from their parents.

## RESULTS

Descriptive statistics are summarized in Table 2. A total of 166 patients were enrolled in the study. Eight patients died before the initial interview, and an additional 18 patients were lost to follow-up or withdrew during the follow-up period. Consequently, 140 patients were included in the final analysis, comprising 91 mild cases and 49 severe cases. The average age for all patients was 16.6 years, and 12.1% were younger than 5 years. The average duration of illness in severe cases was 88.1 days, which was substantially longer than the 22.4 days in mild cases. A total of 8.8% of mild cases were hospitalized, while 95.9% of severe cases were hospitalized. All patients except one had caretakers whereas only four patients had substitute laborers during their illness. The average household income among mild cases was higher than that of severe cases, and a greater proportion of severe cases reported having to borrow money due to typhoid fever.

**Table 2.** Descriptive statistics.

| Item | Mild (n=91) |  | Severe (n=49) |  |
| --- | --- | --- | --- | --- |
| Age, mean (SD) | 16.4 | (12.9) | 17.0 | (13.4) |
| Male, n (%) | 46 | (50.5) | 30 | (61.2) |
| Sick days, mean (SD) | 22.4 | (13.1) | 88.1 | (32.6) |
| Monthly household income, mean (SD), US\$ | 145.3 | (114.0) | 110.8 | (71.6) |
| Proportion of patients studying, n (%) | 60 | (65.9) | 33 | (67.3) |
| Proportion of patients with wage loss, n (%) | 9 | (9.9) | 10 | (20.4) |
| Proportion of patients hospitalized*, n (%) | 8 | (8.8) | 47 | (95.9) |
| Proportion of patients with carers, n (%) | 90 | (98.9) | 49 | (100.0) |
| Proportion of patients with substitute labor, n (%) | 2 | (2.2) | 2 | (4.0) |
| Proportion of patients with borrowed money, n (%) | 29 | (31.9) | 34 | (69.4) |

\* Hospitalized was defined as patients who were interviewed at the hospital while admitted at least once during the interview period

The age distribution of both mild and severe cases was right-skewed, indicating that most of the patients were under 20 years of age (Figure 1). However, relatively few severe cases occurred among children under 5 years of age.

**Figure 1.**
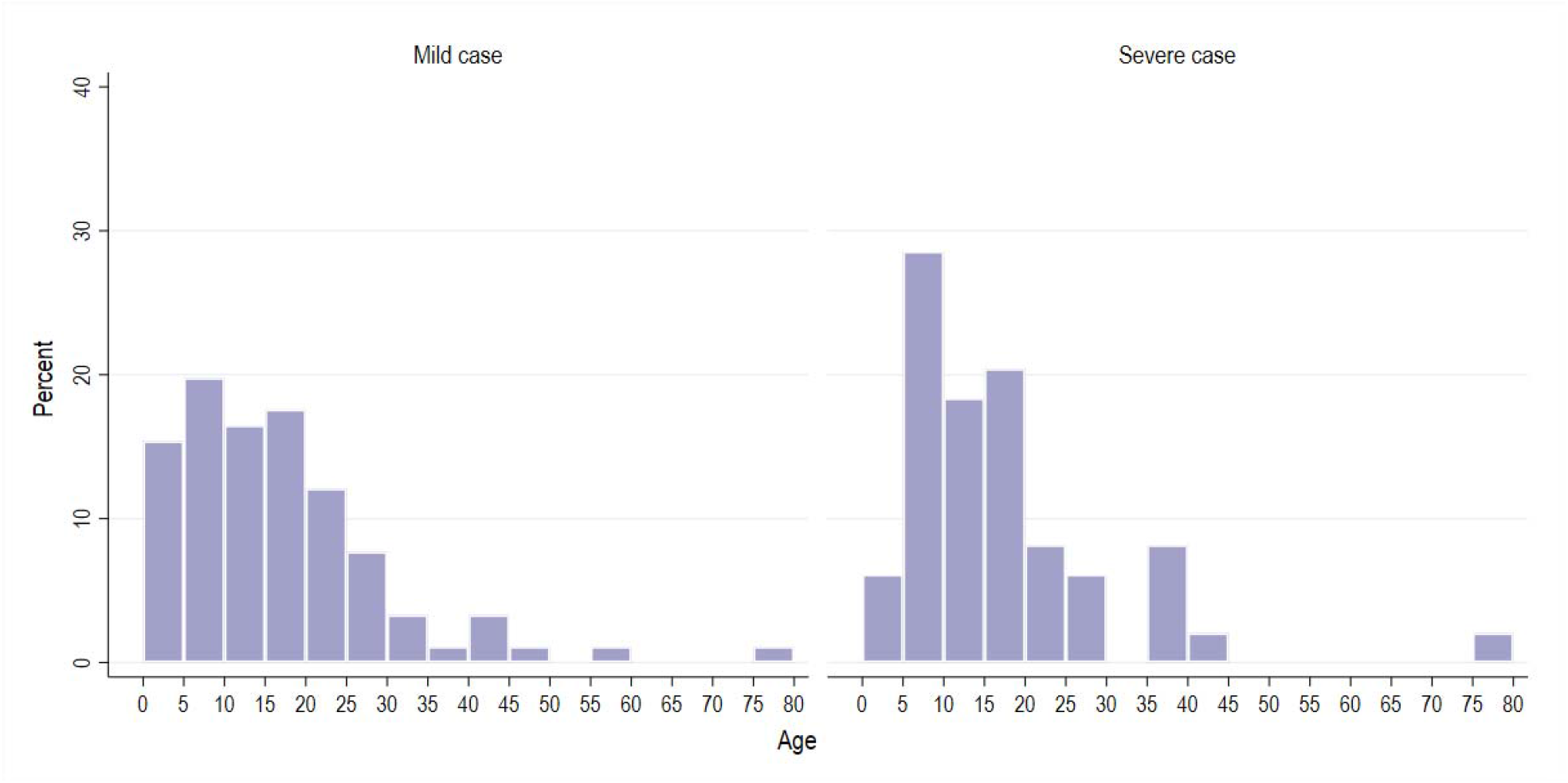
Age distribution.

Among mild cases, public health facilities were the most frequently visited (42.1%), followed by pharmacies (33.2%) and private hospitals/clinics (22.3%) (Figure 2). For severe cases, private hospitals/clinics were the most visited (56.6%), followed by public health facilities (23.4%).

**Figure 2.**
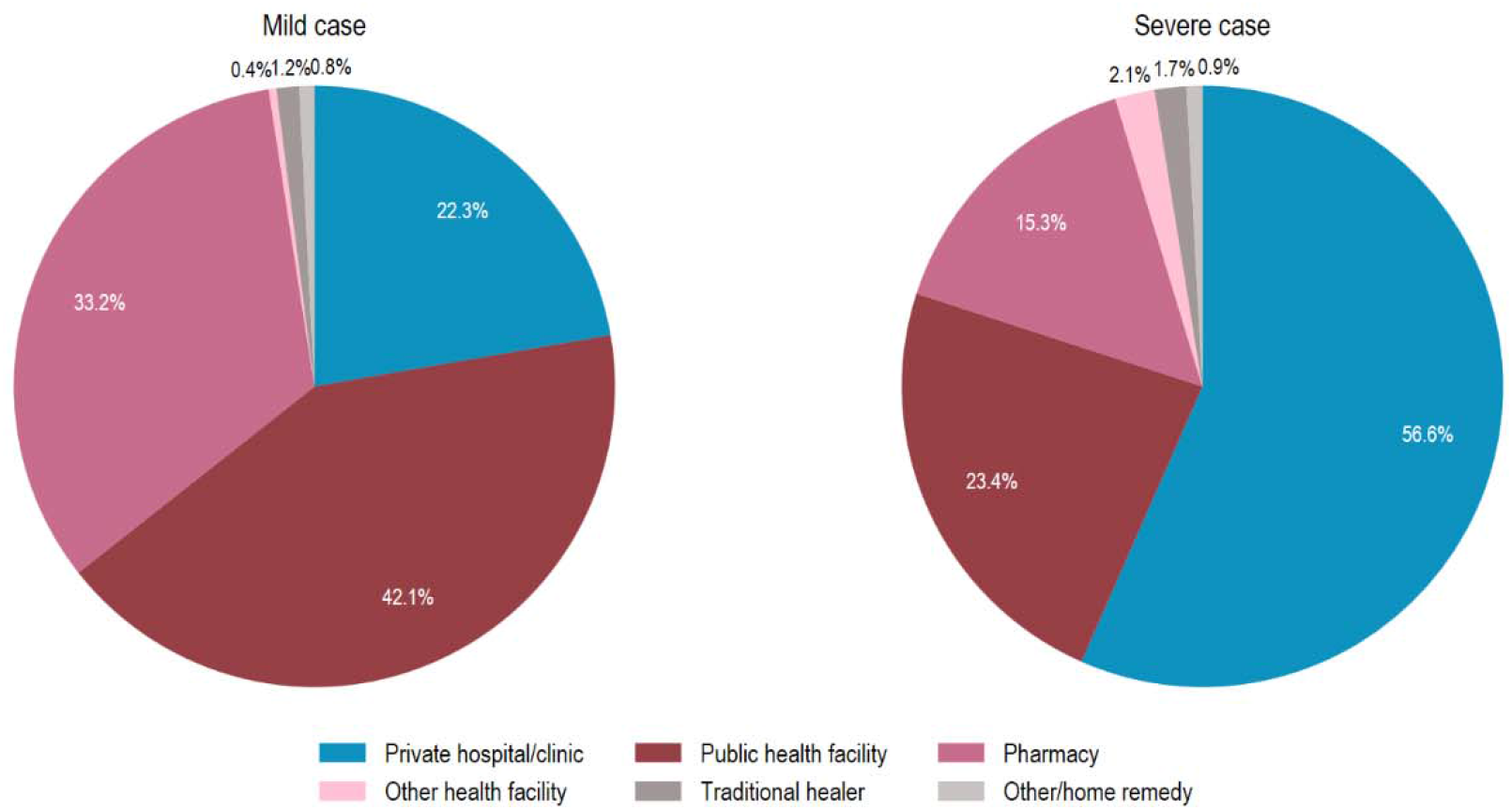
Facility type visited by patients.

At the initial interview, 92.8% of severe cases reported limitations in usual activities due to typhoid fever; of these, 54.1% were unable to perform usual activities (Figure 3). Among mild cases, although the proportion was less than in severe cases, 69.6% reported that their usual activities were affected by typhoid fever. The proportion of patients reporting limitations in usual activities decreased over time. During the last follow-up interview period, 15.9% of severe cases and 8.1% of mild cases reported that they were still unable to perform usual activities.

**Figure 3.**
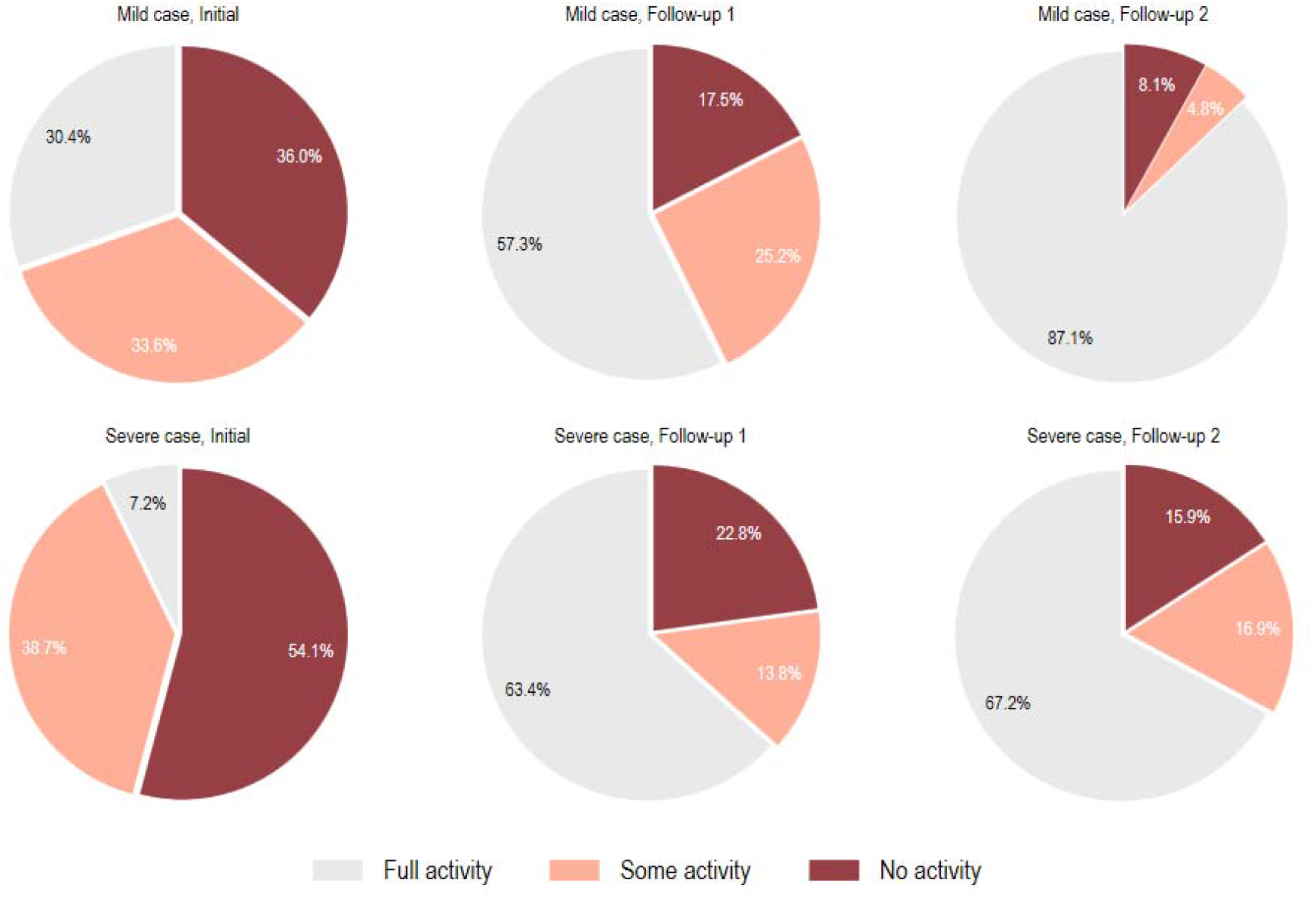
Activity level by interview period.

The mean economic burden per typhoid fever episode was estimated at US$ 273 (Figure 4). Specifically, US$ 579 (95% CI: US$ 506–663) for severe cases and US$ 108 (95% CI: US$ 90– 127) for mild cases. Median costs (interquartile ranges) were US$84 (US$40–151) for mild cases and US$512 (US$403–688), indicating a right-skewed distribution of costs in both groups. Among the three main cost components, DMC represented the largest cost component in severe cases, whereas IC represented the largest cost component among mild cases. None of the participants were covered by insurance or hospital subsidies, indicating that all DMC were paid out-of-pocket by households. Productivity losses associated with premature mortality were also estimated. During the study period, eleven deaths were observed. The mean productivity loss per premature death was estimated at US$15,987. Eight patients who died before the initial interview were excluded from the episode-based COI analysis.

**Figure 4.**
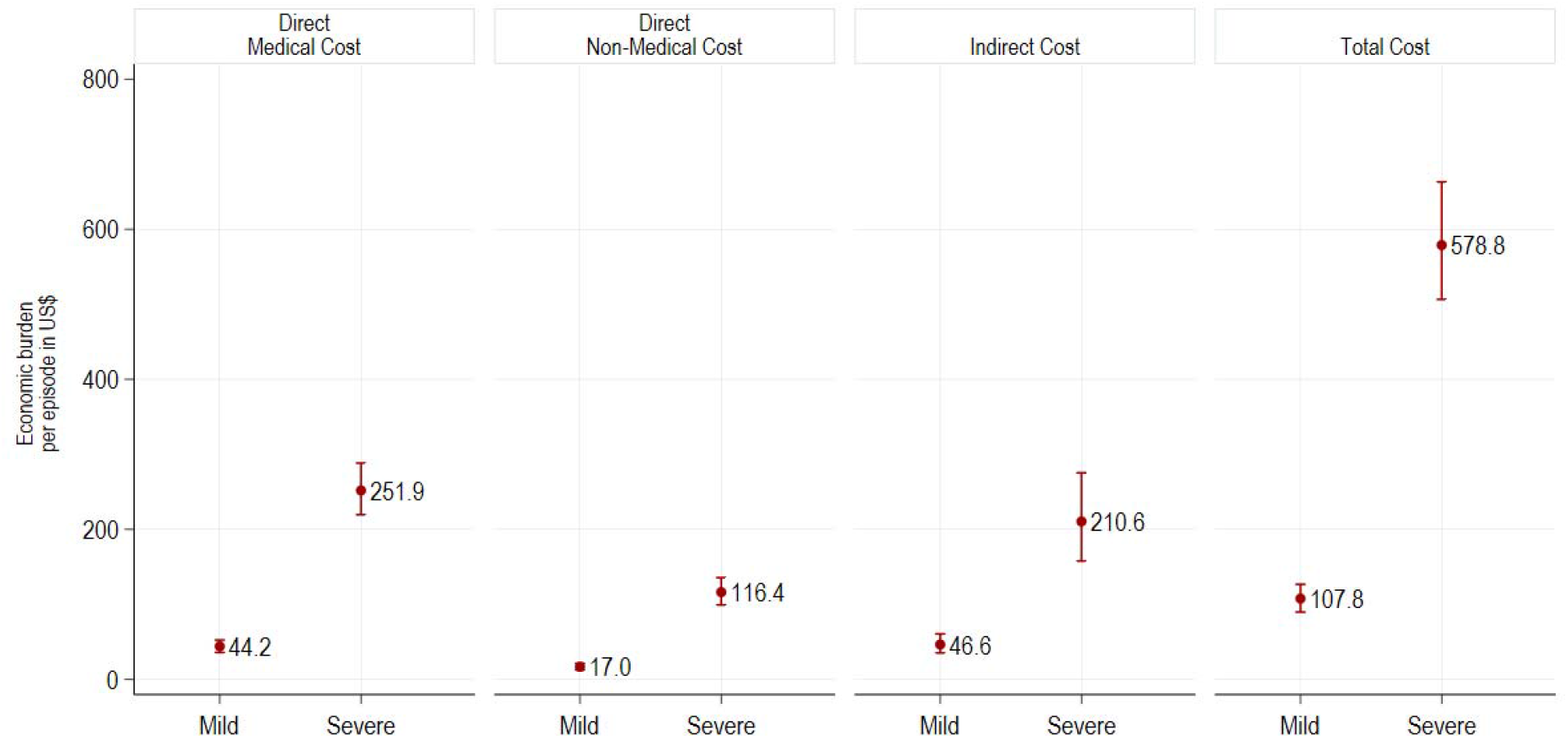
Cost of Illness.

In both mild and severe cases, the average burden was highest in the 5–14 age group, followed by those aged 15 and over, and lowest among children under 5 years (Figure 5). However, in severe cases, a wider confidence interval was observed in those under 5 years, probably reflecting the small number of severe cases in this age group.

**Figure 5.**
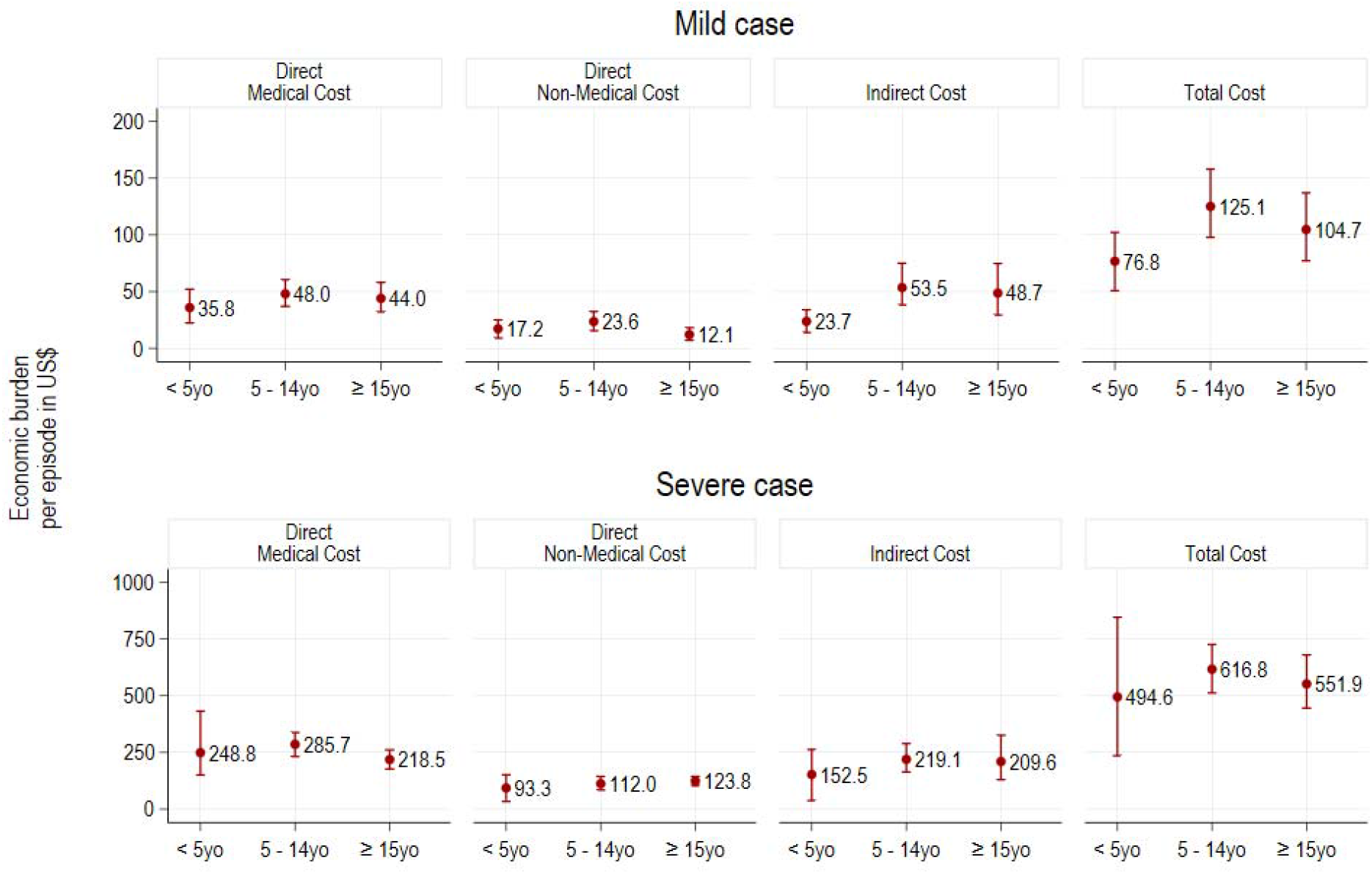
Cost of Illness by age group.

Among the 132 participants with available household income data, 79 households (59.8%) incurred economic burden due to typhoid that exceeded their monthly household income (Figure 6). Of these, 36 were mild cases and 43 were severe cases. A larger proportion of severe cases experienced costs exceeding monthly household income.

**Figure 6.**
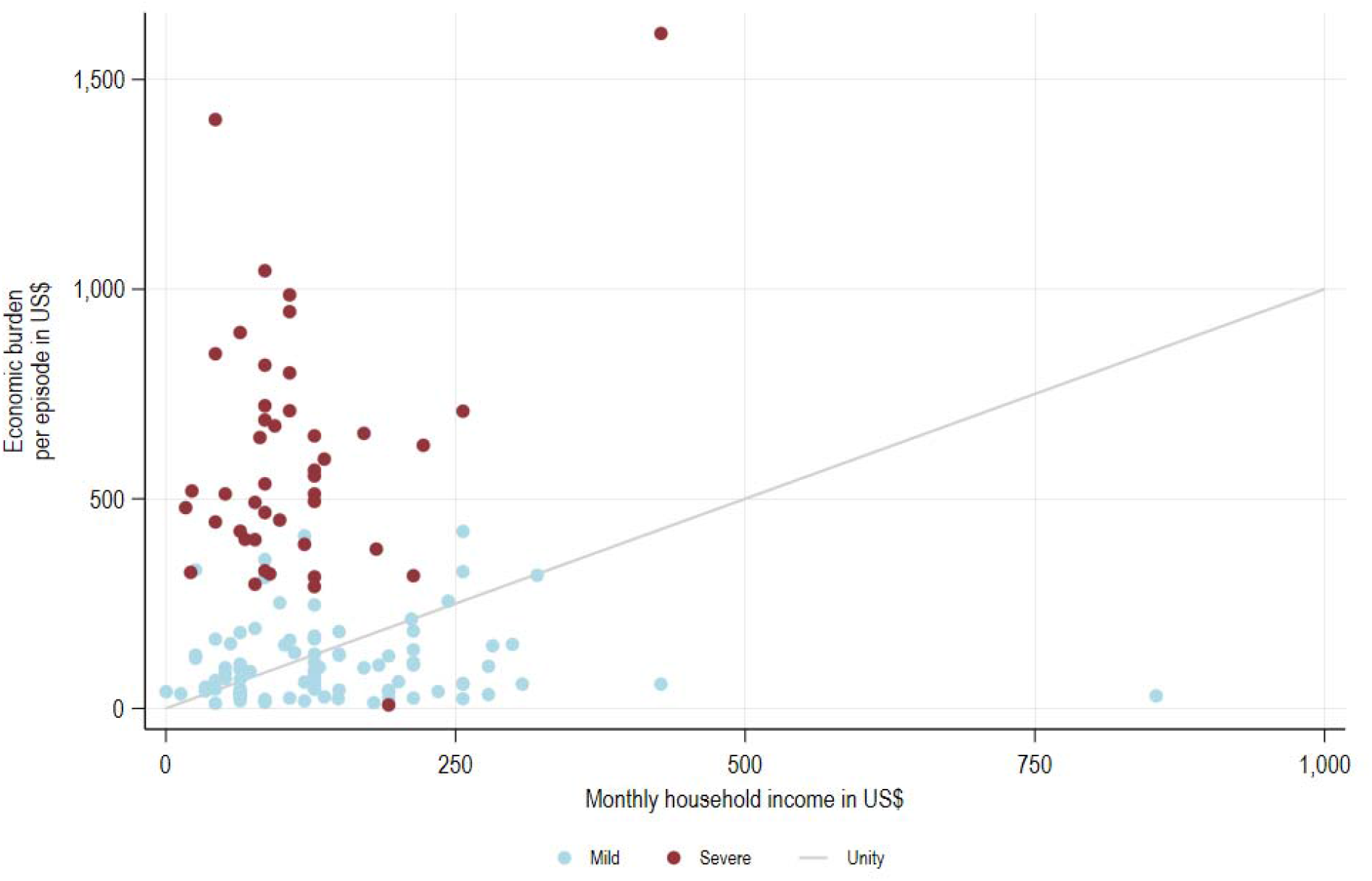
COI vs monthly household income.

## DISCUSSION

COI estimates for typhoid fever were generated using patient-level survey data collected in the Kisantu Health Zone. No previous empirical COI study from the DRC was identified in the recent literature review (7), making these estimates the first primary evidence available for the country.

This study estimated the economic burden of typhoid fever at US$ 108 for mild cases and US$ 579 for severe typhoid cases with suspected intestinal perforation or peritonitis. In the 13 studies reviewed by Debellut et al. (7), reported outpatient costs varied between US$20 in India and US$143 in China, whereas estimates for ileal perforation ranged from US$551 in Niger to US$1,735 in India. Because approximately 91% of mild cases were managed without hospitalization, the estimated cost for mild disease was close to the upper end of previously reported outpatient estimates, while the estimate for severe disease remained within the reported range for ileal perforation. Indirect costs represented 43.2% of the total cost, exceeding the proportions reported in previous studies.

The economic burden of typhoid fever corresponded to approximately two months of average household income, consistent with findings reported from Tanzania (20). For severe cases, the economic burden was estimated to reach nearly five times the monthly household income. Given that none of the patients in this study were covered by insurance, the financial burden associated with healthcare expenditure was likely considerable. In total, 45% of participants reported that they had to borrow money due to typhoid fever, indicating that typhoid fever was associated with substantial expenses and income losses, resulting in considerable financial hardship for many households.

The study had several limitations. Patients who signed the consent form but passed away before the initial interview, or patients who were unable to complete follow-up interviews for reasons such as relocation, were excluded from the COI study. Because these patients died before any cost information could be collected, the healthcare costs associated with terminal care could not be estimated. The overall economic burden of severe typhoid fever may have been underestimated. Although follow-up interviews were conducted over approximately 100 days, longer-term healthcare costs and productivity losses beyond the follow-up period may not have been fully captured. In addition, severe cases were defined using clinical criteria without systematic microbiological confirmation. Therefore, some cases may have been misclassified because other acute abdominal conditions can present with similar clinical features. Productivity losses among students were valued using government expenditure per primary school student as a proxy. However, this proxy may not fully represent the actual economic value of students’ time lost because of illness and school absenteeism, and these estimates should be interpreted with caution.

This study estimated patient-level costs incurred by patients and their households; healthcare provider costs were not included. Consequently, the estimated costs do not reflect the full economic burden of typhoid fever treatment. In addition, household expenditures were based on patient-reported information and may have been affected by recall bias, potentially resulting in either underestimation or overestimation of reported costs. Furthermore, because the Kisantu Health Zone operates under a flat-fee system to improve access to quality health care (21), patient-reported expenditures may not fully reflect the underlying cost of healthcare services. Because the study was conducted in a single health zone, caution is needed when generalizing these findings to other regions of the DRC with different socioeconomic, geographic, and healthcare system characteristics. Further research incorporating provider-side cost estimates from multiple settings would provide a more comprehensive assessment of the economic burden of typhoid fever.

The findings indicate that typhoid fever places a considerable financial burden on affected households in the DRC. As the WHO has recommended the use of TCVs in countries where typhoid is endemic (8), these findings provide important evidence to inform TCV introduction and other typhoid control policies. Beyond disease prevention, the substantial out-of-pocket burden observed in this study highlights the need to strengthen financial protection mechanisms.

Although a flat-fee system is implemented in the Kisantu Health Zone, households may still face considerable treatment-related expenditures, particularly in severe cases. Expanding prepayment and risk-pooling mechanisms, increasing public or external subsidies, and reducing direct payments at the point of care may help lower the risk of catastrophic health expenditure. This study contributes important empirical evidence on the economic burden of typhoid fever in LMIC settings, where such data remain limited.

## Data Availability

Aggregated data may be made available by the corresponding author upon reasonable request and subject to the conditions of the ethical approval.

## Acknowledgements

We thank the members of the An open-label effectiveness study of a typhoid conjugate vaccine in Kisantu, Democratic Republic of the Congo team for their contributions and support. We also would like to acknowledge generous funding provided by the European & Developing Countries Clinical Trials Partnership, Bill & Melinda Gates Foundation.

The International Vaccine Institute gratefully acknowledges the partnership and financial contributions provided by the governments of the Republic of Korea, Sweden, Austria, Finland, India, Thailand, and the Philippines.

